# Pilot proof for RNA biomarker-based minimally invasive endometrial receptivity testing using uterine fluid extracellular vesicles

**DOI:** 10.1101/2023.03.02.23286679

**Authors:** Alvin Meltsov, Elisa Giacomini, Paola Vigano, Natasa Zarovni, Andres Salumets, Elina Aleksejeva

## Abstract

**Research question:** How accurate is minimally invasive assessment of endometrial receptivity from uterine fluid extracellular vesicles (UF-EVs) using only 68 RNA biomarkers.

**Study design:** Assessment of endometrial receptivity by applying the beREADY computational model on transcriptomic data derived from UF-EVs during the pre-receptive (LH+2) and receptive phases (LH+7) of a natural cycle.

**Results:** beREADY housekeeping gene transcript levels in UF-EV are not significantly different between LH+2 and LH+7 UF-EVs. Endometrial receptivity can be assessed from UF-EV transcriptomic data using beREADY computational model with specificity of 100% and sensitivity of 75%.

**Conclusion:** beREADY computational model can be used on UF-EVs instead of endometrial tissue biopsy to assess endometrial receptivity. However, further development is needed to fine-tune the model to show that this model can also be used on UF-EV samples from women undergoing hormone-replacement therapy.

## Introduction

beREADY test has been originally developed to assess endometrial receptivity from a tissue biopsy using 68 transcript biomarkers and 4 housekeeping genes (1). However, the biopsy procedure excludes the possibility of embryo transfer (ET) in the same cycle and thereby prolongs IVF treatment. Furthermore, current data regarding the efficacy of endometrial receptivity assessment is unclear. One of the factors affecting implantation success irrespective of receptivity testing could be cycle-to-cycle variation of window of implantation. Current methods are invasive, requiring a tissue biopsy from the endometrium - as a result, receptivity testing is done during cycle preceding ET. Therefore, receptivity assessment right before ET would likely improve predictive value of the receptivity testing as possible cycle-to-cycle variations in the window of implantation would be circumvented. Recently, endometrium shed extracellular vesicles (EVs) in uterine fluid (UF) have been described as carriers of tissue biomarkers for non-invasive assessment of endometrium status and dynamics (2). UF aspiration before ET is minimally invasive and available data shows it is not detrimental to the implantation rate (3,4). Consequently, to develop a minimally-invasive receptivity assessment method, in this study beREADY computational model has been applied on the transcripts contained within EVs that were isolated from UF (2) (Figure 1A).

**Figure 1.**
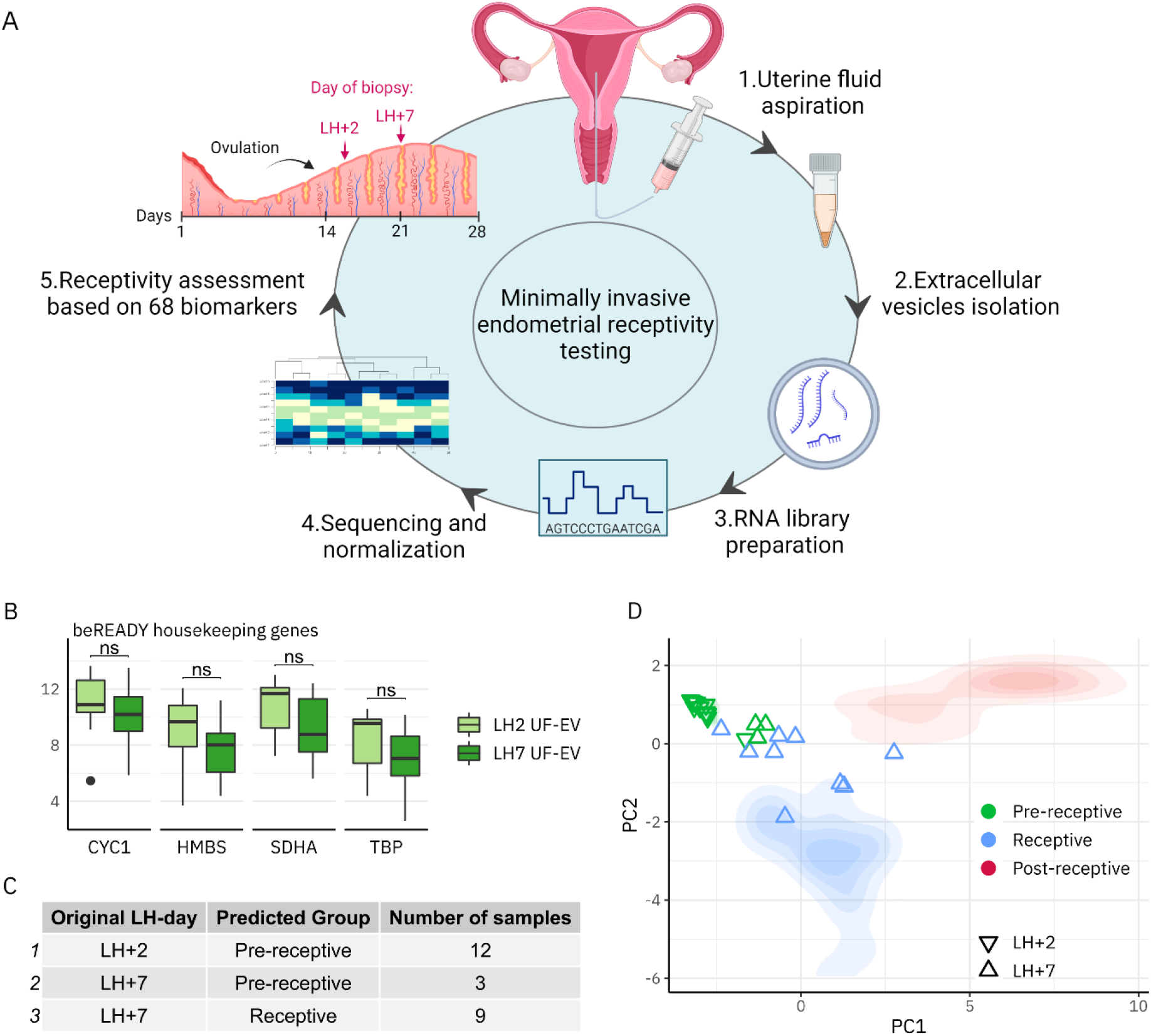
Application of beREADY model of endometrial receptivity assessment on UF-EV samples. **A)** Graphical representation of minimally invasive beREADY test. **B)** The detection of beREADY housekeeping gene transcripts in UF-EV transcriptomes. Two-sided t-testing shows non-significant expressional differences in beREADY test results when comparing LH+2 with LH+7 UV-EV samples. **C)** beREADY computational model assessments of receptivity based on UF-EV transcriptomes. **D)** Principal component analysis (PCA) plot shows the distribution of analysed LH+2 and LH+7 samples (nablas and triangles) among beREADY reference groups (two-dimensional visualization of distribution in the PCA space). beREADY reference groups (pre-receptive, receptive, post-receptive) are based on receptivity assessments of endometrial tissue biopsies done with histological evaluation of biopsies according to Noyes’ criteria.

## Methods

### Study group and sample collection

The study group consists of fertile women - their selection is described in (2). Uterine fluid samples from LH+2 (n=12) and LH+7 (n=12) of the natural cycle were collected as described in (2).

### EVs isolation and sequencing library preparation

EVs were isolated by differential centrifugation as described in (2). Sequencing libraries were prepared using SMART-Seq v4 Ultra Low Input RNA Kit (Takara Bio Inc.) as described in (2).

### Data analysis

beREADY computational model applied in this work was developed as described in (1).

## Results

### beREADY housekeeping genes can be applied for the normalisation of UF-EV samples

To assess whether beREADY housekeeping genes (*CYC1, HMBS, SDHA, TBP*) are consistently detected in UF-EV transcriptome and are stable across different menstrual cycle phases, their levels were analysed in LH+2 and LH+7 UF-EV samples collected from fertile women (Figure 1B). beREADY housekeeping genes were detected in all samples and there was no significant difference in the quantity of these transcripts between LH+2 and LH+7 confirming their usability as housekeeping genes. Noteworthy, in the same sample set, the expression of EV candidate housekeeping genes (5) was also analysed and no significant changes were found between LH+2 and LH+7 (data not shown).

### beREADY model classifies LH+2 and LH+7 UF-EV samples based on receptivity status

Next it was inquired whether the beREADY model correctly classifies LH+2 (pre-receptive, n=12) and LH+7 (receptive, n=12) samples from naturally cycling fertile women. All LH+2 samples were correctly classified as pre-receptive, whereas only 9 of the LH+7 samples were correctly classified as receptive (Figure 1C,D). Based on this data, the beREADY model sensitivity is 75%, and specificity is 100%.

## Discussion

This pilot study shows that beREADY computational model can be used to assess endometrial receptivity in a minimally invasive manner from UF-EV transcriptomic data with sensitivity of 75% and specificity of 100%. The lower sensitivity is expected since the beREADY computational model was developed for tissue biopsies in conjunction with a targeted sequencing assay and not the whole transcriptomic data used in this study (1,4). Alternatively, it cannot be ruled out that the endometrial maturation in these three LH+7 women classified as pre-receptive was slower and corresponded to the pre-receptive status. Future studies consisting of parallel analysis of endometrial tissue and UF-EV samples are necessary. Furthermore, it is not known whether the model is accurate for assessing receptivity during hormone-replacement therapy. A minimally invasive endometrial receptivity test would allow to assess endometrium right before ET and therefore minimise diagnostic errors due to possible window of implantation shifts from one cycle to another.

## Data Availability

Raw data has been already published.

